# Correlating Covid-19 mortality and infection levels

**DOI:** 10.1101/2020.05.01.20087320

**Authors:** Mugdha Gadgil, Chetan Gadgil

**Affiliations:** Chemical Engineering and Process Development Division, National Chemical Laboratory, Dr Homi Bhabha Road, Pune 411008, India

## Abstract

Covid-19 deaths and positive cases show a remarkable heterogeneity across countries which cannot be easily explained on the basis of similarities or differences in the quality of healthcare, access to healthcare, testing facilities, or preventive measures such as lockdowns. Here we show that there is a distinct correlation between the mortality level and the infection level across countries, which can explain the mortality levels for a wide spectrum of countries. This implies that the number of deaths per 100 infected individuals is approximately the same across diverse countries and can be estimated from the slope of the mortality level-infection level plot. The correlation presented here can potentially be combined with estimates of infection spread to forecast future mortality levels and therefore future needs in terms of healthcare and other resources. Tracking of an individual location’s temporal path on this plot can potentially serve as a visual assessment of the nature of the epidemic. Methods presented here are not specific to the current epidemic. This is a preliminary report and uses data from a single source at a single time-point to demonstrate the capability of such an analysis.

## Introduction

Estimating the requirement of hospital resources is critical to managing the ongoing Covid-19 epidemic. The number of deaths is likely to be a good basis for estimating the number of hospital beds required. Here we attempt to estimate the expected mortality level from a given infection level. We show that the data is remarkably consistent across countries, and therefore believe that these results will be relevant across geographical locations.

### Hypothesis

There is a large variation in deaths per million individuals across countries, that seem to be unexplainable on the basis of amount of testing, sophistication of the healthcare system, and whether lockdown measures were employed. We hypothesize that the mortality level as a fraction of infected individuals is constant, and the variation can be explained primarily on the basis of difference in infection level.

We assume that if a sufficiently large number of tests (relative to population size) are conducted, the infection level calculated from these tests is a reasonable estimate of population infection level. Strictly speaking, it is an upper bound on the infection level, since it is more likely that infected persons will get tested.

Once the infection level and the relationship between mortality and infection level is estimated, the mortality can be calculated and resources scaled by this number can be allocated accordingly. Alternately, when testing is inadequate, based on known mortality, the current level of infection in the population can be estimated in the absence of random population-level testing.

## Methods

We used publicly available data^1^ on total (positive) cases, total deaths, total tests, and total deaths per million population for 214 countries. We do not consider countries that have fewer than 100 deaths, or have tested at a level less than 7000 tests per million population. For the 27 remaining countries, we estimate infection level as the total number of cases divided by the total number of tests. The cutoff numbers (100 deaths and 7000 tests per million) are arbitrary, so we have repeated the analysis for 9 different combinations of minimum deaths (75, 100, 125) and minimum testing level per million population (5000, 7000, 9000).

The linear fit to these numbers is visually skewed by the presence of a number of data points at low infection levels, and sparser points at higher infection levels. To check whether the linear fit holds for low infection levels we consider only those countries from the 27 that have low (<7.5%) infection levels. We find that the linear fit is still valid in this range, but is clearly not correct for infection levels below 3%: for instance, at infection level= 0, it predicts a negative value for mortality. If there is a pressing requirement to use the fit at lower infection levels despite the caveats stated here, we have plotted a power-law and exponential fit, both of which will result in positive mortality predictions irrespective of infection level, and have a R^2^ better than the linear fit. We emphasize that none of the equations is intended to suggest either a mechanism or a trend beyond the range of infection levels in that figure. Prudence (and Occam’s razor) suggests that when there is a choice, a linear fit is to be used.

## Key Result: Mortality level is a linear function of infection level

Figure 1 shows the graph of deaths per million (henceforth referred to as mortality level) as a function of estimated infection level (calculated as total positive cases/total tests). The data seems to indicate that the relationship between the mortality level and infection level can be approximated by a linear fit. Countries with similar socioeconomic conditions differ in mortality primarily due to a difference in infection levels, at least in the 3% to 25% range. There are eight clear outliers, three with a higher than predicted mortality and five with a lower than predicted mortality.

**Figure 1:**
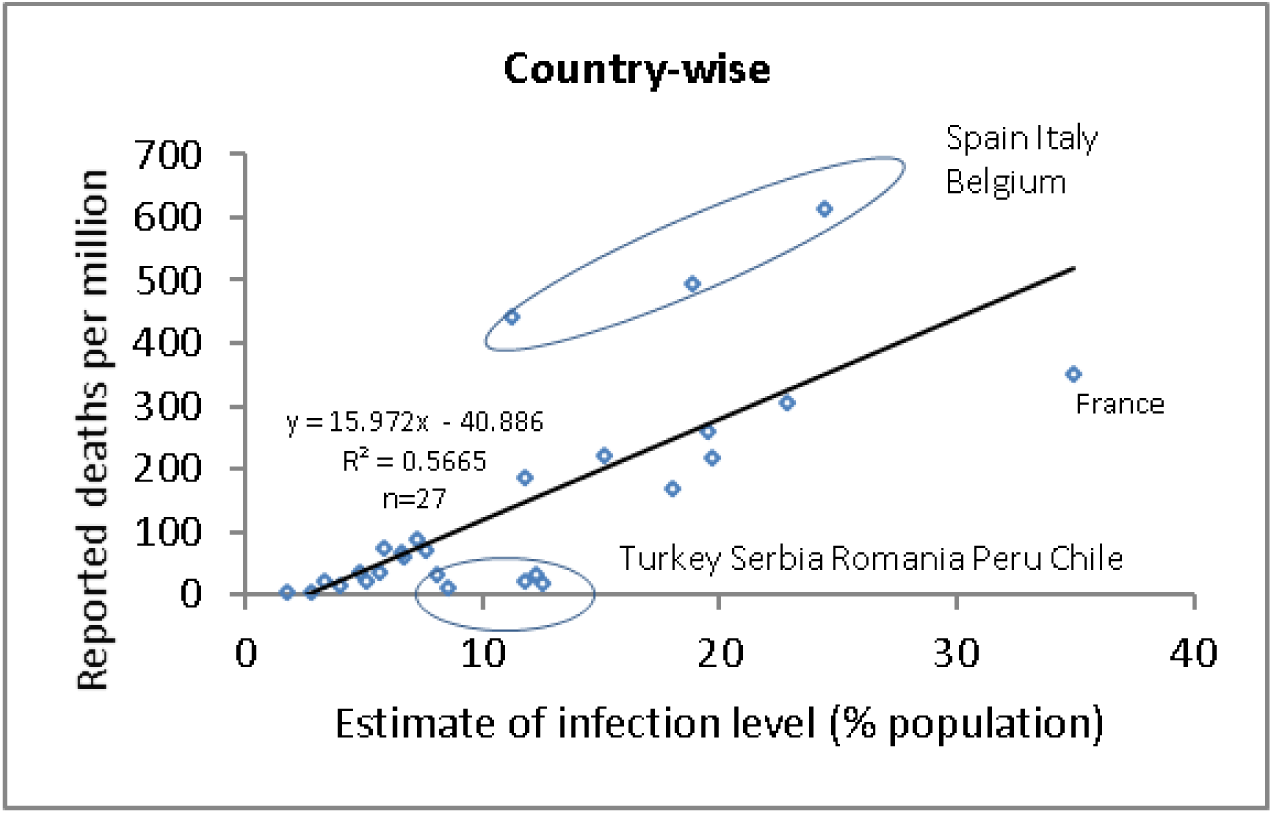
Mortality-infection correlation.

We have tested the effect of removing all eight countries from our analysis (Figure 2). As expected, the fit is better. We note that such an ‘arbitrary’ removal of data is not statistically valid. However, this is not a standard statistical data set, since each point represents a different country with a different healthcare system, political system, reporting system, etc. Figure 2 shows that despite these differences, 19 countries show a clear linear trend relating mortality level to infection level.

**Figure 2:**
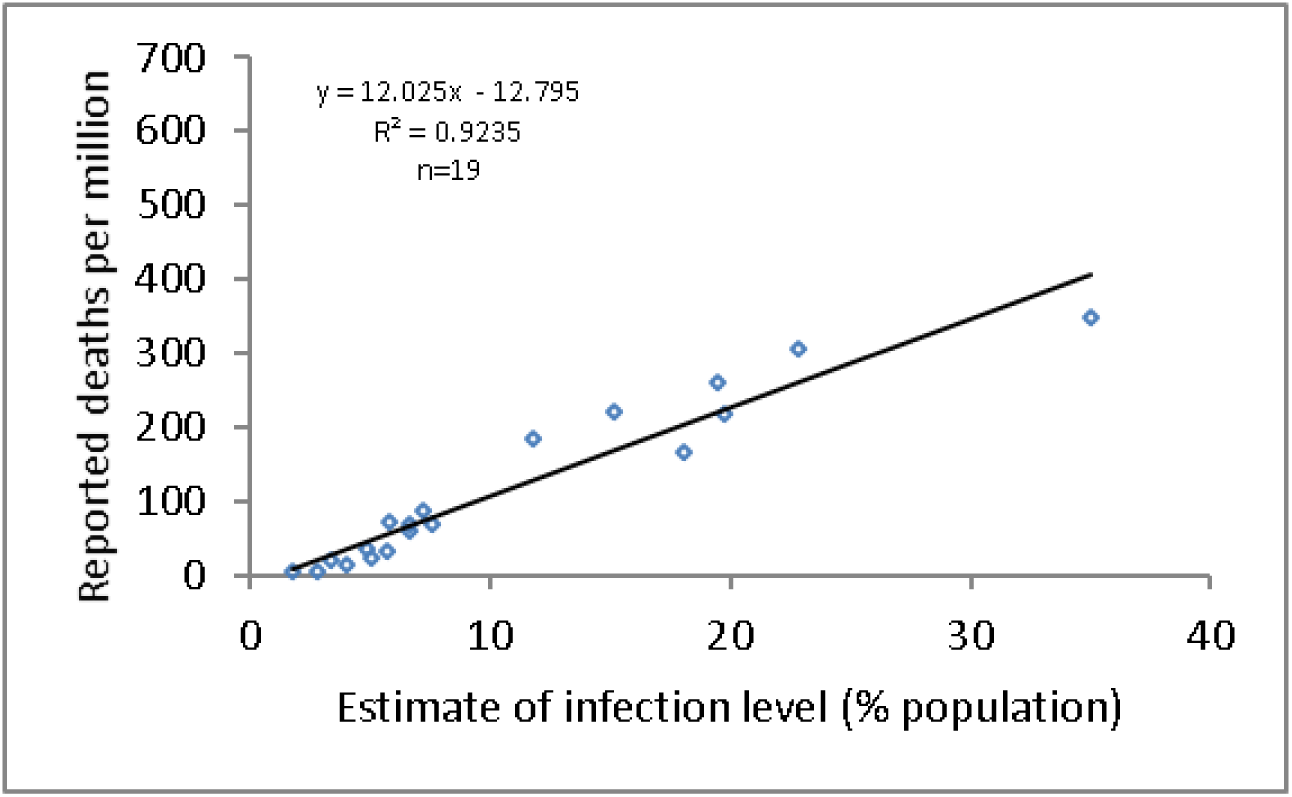
Figure 1 with 8 countries removed from analysis.

We next explore the trend at low infection levels, which are currently estimated to be prevalent in India from the ratio of positive tests to total tests. Figure 3 shows the data in Figure 1, limited to infection levels < 7.5%. We can use the data to estimate (an upper bound for) infection level. For instance, the dashed purple horizontal line represents the reported mortality level for Pune, which is ~10 with reported ~68 deaths and a population of 7 million^3^. For this mortality level, the infection level is predicted by all data fits to be slightly less than 3%. We reiterate that the use of power law and exponential fits is only intended to constrain estimates for low infection levels to be positive, and are not meant to suggest either the nature of the mortality-infection level relationship, or any mechanism. Applying power law and exponential fits to the whole data in Figure 2 or 3 does not result in significant improvement to the fit.

**Figure 3:**
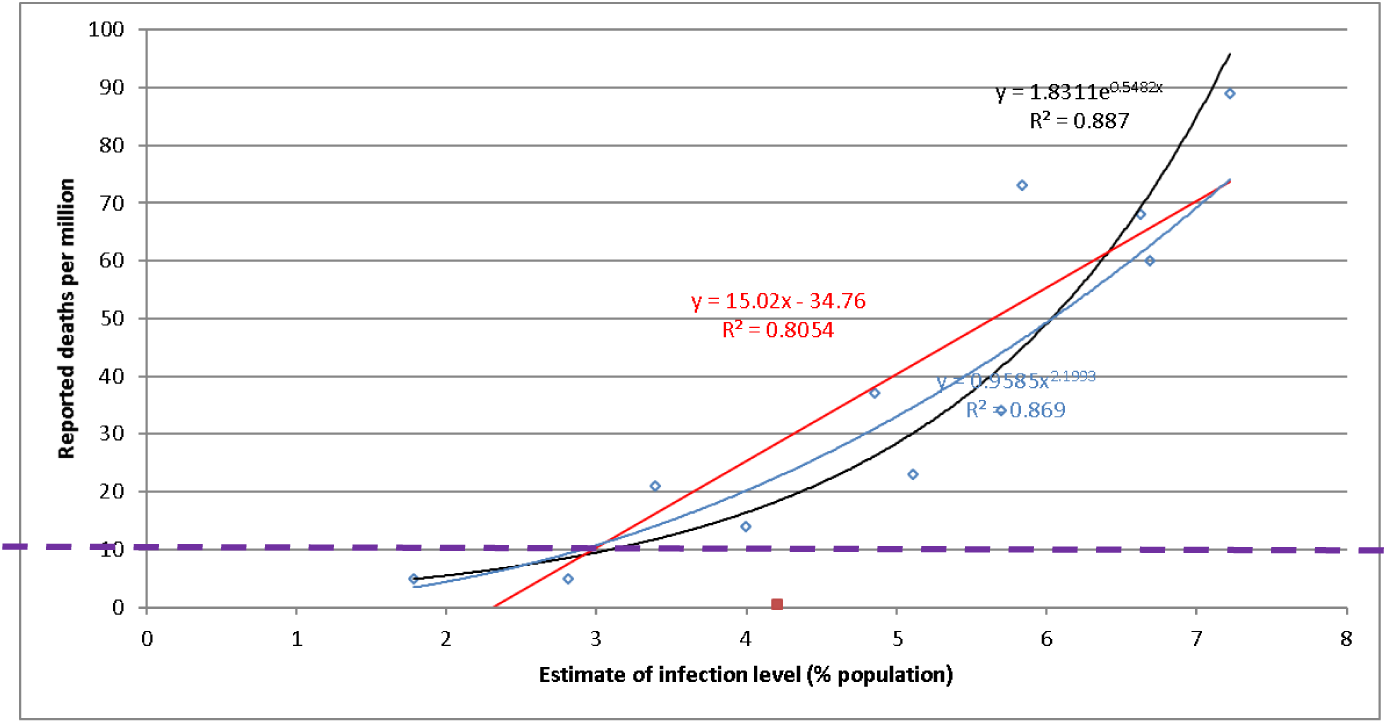
Countries with low infection levels.

For all of the curves, the slope of the line (or derivative of the function) scaled appropriately^4^ will give an estimate of the fraction of infections that result in death. Since the relationship appears to be linear, we conclude that this number is constant across countries and across population infection levels. This may mean that there is a single strain, or multiple ones with similar macro-characteristics; and therefore, the experience of other countries may serve as a good guide to estimate requirements. From Figure 2, we estimate this number to be 0.12 (Figure 2) or 0.15 deaths/100 infected persons (Figure 1, Figure 3).

We check to see how well the predictions fit data that is not used in constructing the model. Figure 4 uses data from the same website but for individual US states, applying the same filters for testing and deaths as data used in Figure 1. The black line is the prediction copied from Figure 2, and the red line is the linear fit just to US state-wise data. This exercise captures the uncertainty in applying this analysis to other countries (such as to India), and supports a limited predictive capability of the analysis.

**Figure 4:**
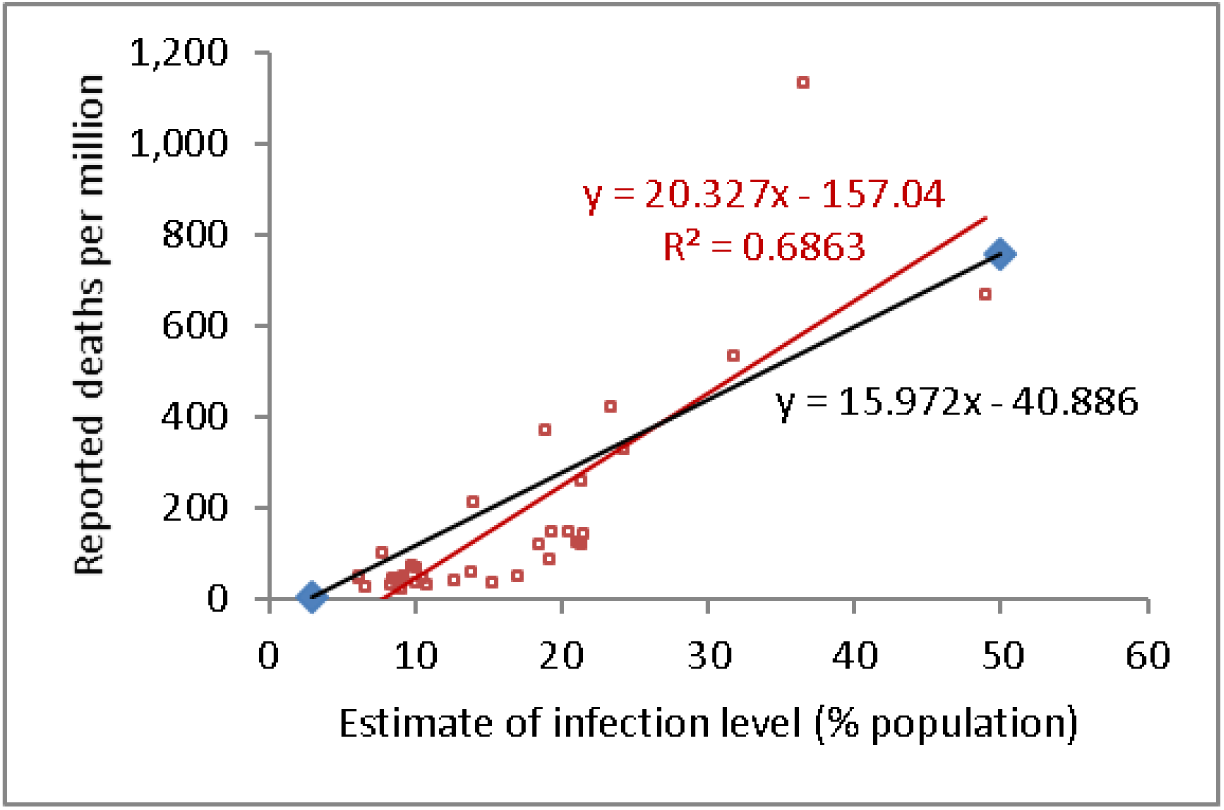
Prediction and actual values for individual US states.

Finally, we test the effect of varying the cutoffs applied for scale of testing and the number of deaths (see methods). Supplementary Figure 1 shows the mortality-infection plot in Figure 1 with differing cutoffs for minimum testing level (5000, 7000, 9000 tests per million) and minimum number of reported deaths (75, 100, 125). Even with these values, the number of data points (countries exceeding cutoff criteria) increase or decrease by a maximum of 7 from the 27 countries in Figure 1. There is no perceptible change in the nature of the data trend. The figures in the last row (cutoff of 125 deaths) are the same as the corresponding figures in the center row (cutoff of 100 deaths as for Fig 1). Increasing the stringency to 9000 tests per million and 125 deaths results in the elimination of four countries that had lower mortality level than predicted by the linear correlation in Figure 1. This results in higher slope of 0.19, i.e. different from the 0.12-0.15 slopes in Figures 1 and 2.

## Discussion

We have used publicly available data to show that there is a clear relationship between infection level and mortality level. This relationship and plot can be used in several ways:

1. Estimating an upper bound for infection level from reported deaths, as shown for Pune (Fig 3).
2. Estimating healthcare infrastructure requirements: From an estimate of the infection level using epidemiology models or other data, future mortality levels can be estimated from the correlations in this report and the corresponding mortality number and bed requirement can be calculated. For instance, at a 20% infection level, the linear correlation predicts 227 deaths/million (from Fig 2) or 279 deaths/million (from Fig 1). This correlation implies that if the prediction from an epidemiological model is that a 20% infection level (similar to current levels for Sweden, UK, Netherlands) will be reached in Pune (7 million population) in three months, the linear correlation (Fig 1,2) predicts that there will be ~1500 deaths in the next three months. South Korea which has reported zero new cases in the past few days seems to have limited the infection level. If Pune limits spread to say 10%, the linear correlation predicts a mortality level of 118 deaths/million (Fig1) or 107 deaths/million (Fig2), which corresponds to ~750 deaths in Pune for this assumed 10% infection level.
3. It is estimated that herd immunity will occur (i.e. new cases will reduce to almost zero) when a certain fraction (that depends on the reproduction number R0) is infected. For R0 close to 3 this level would be ~70%. The data does not extend to those infection levels, and therefore the urge to extrapolate should be vigorously resisted.
4. This analysis presents a means of visually assessing the time-course of infections. It will be very informative to plot a time-course for every country on such infection level-mortality level graph. A linear plot will signify lack of mutation or lack of stress on hospital resources. Nonlinearity may be indicative of changing strains leading to differing mortality per 100 infected individuals, or a change in circumstances such as overburdening of hospital resources or an infusion of resources that lead to changed mortality per 100 infected individuals with time for a particular country or location. Differences in slope between locations may arise due to differing susceptibility arising from a difference in demographics, especially for diseases that are known to disproportionately affect particular age groups. We believe that an analysis of epidemic types can be facilitated from such curves on the mortality-infection plot.

There are several caveats about the assumptions and therefore predictive ability of the model *even if the data is extremely accurate*. The accuracy of the reported numbers is unknown and is expected to vary across countries. We have assumed a linear model since it is the simplest possible fitting function. This choice however does imply any mechanistic basis. All estimates for mortality numbers critically depend on the validity of the estimate for infection level in addition to the uncertainty inherent in the correlation presented here. With the data available, it is not possible to discount the possibility of the correlation in Figure 1 or Figure 2 not applying to some countries/locations due to differences in population susceptibility arising from factors such as population demographics.

Given these caveats, any estimates should be taken as numbers resulting from back-of-the-envelope calculations which is what this analysis amounts to. Nevertheless, given the apparent consistency across countries, we believe that such an analysis can contribute to the estimates for healthcare capacity planning, and such mortality-infection plots can be an aid to a visual analysis of epidemic progression.

## Data Availability

Data used in this article was obtained from a publicly available website https://www.worldometers.info/coronavirus/

---

**Supplementary Figure 1.**
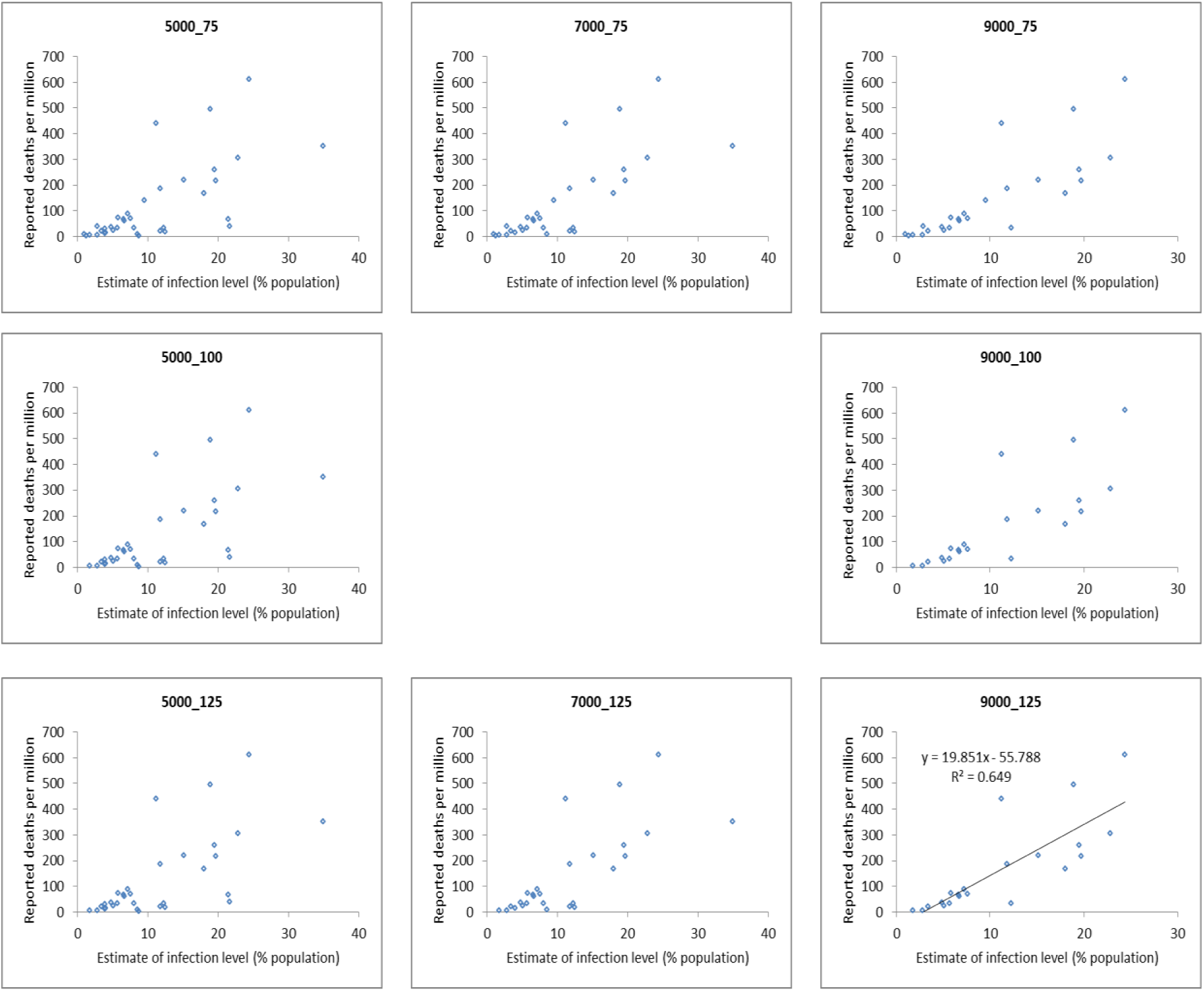
*Mortality-infection plots* for varying parameters of mortality level and infection level used for filtering the data. Chart title shows infection level_ mortality level. Missing central chart is Figure1.

## Footnotes

1 https://www.worldometers.info/coronavirus/ accessed 27 April 2020.

2 Countries in this list are: USA, UK, Turkey, Switzerland, Sweden, Spain, Serbia, S. Korea, Russia, Romania, Portugal, Poland, Peru, Norway, Netherlands, Italy, Israel, Ireland, Germany, France, Finland, Denmark, Czechia, Chile, Canada, Belgium, Austria

3 https://indiapopulation2019.com/population-of-pune-2019.html

4 Slope is deaths per million ÷ infected per 100, So slope/10000 will give or deaths per 100 ÷ infected per 100; and slope/10000*100 (or slope/100) will give deaths per 100 infected.

## Notes

### Competing Interest Statement

The authors have declared no competing interest.

### Funding Statement

No external funding related to this project was received.

